# Early effect of the Covid-19 epidemic on vaccine coverage of major antigens in Guinea: an analysis of the interrupted time series of national immunization coverage

**DOI:** 10.1101/2020.09.11.20192161

**Authors:** Moustapha Dabo, Yombounaud Samah, Mouctar Kande, Djenou Sompare, Aly Camara, Bah Mamadou Dian, Sekou Solano, Idrissa Baldé, LATOU Fabrice, Younoussa Sylla, Iya saidou Condé, Gerard Cristian Kuotu, Almamy Amara Touré, Magassouba Aboubacar Sidiki

**Author notes:** **Correspondence to:** Dr. Magassouba Aboubacar Sidiki, MD: Expanded Immunization Program (EPI), Guinea.

## Abstract

**Introduction:** Since the declaration of the first case of Covid-19 on March 12, 2020, in Guinea, the number of COVID-19 cases has been increasing day by day despite the state of health emergency and the barrier measures decreed by the Guinean government. This present study aimed to assess the early impact of COVID-19 on vaccine activities by comparing current trends to trends over the past year when vaccine coverage of major antigens (BCG, OPV, DTP-HepB-Hib, MMR, IPV, and Td) had improved considerably.

**Methods:** The study was carried out at the Expanded Vaccination Program (EPI) of the Republic of Guinea from February 2020 to June 2020. It was a comparative retrospective cohort study on the trends in administrative coverage of the different antigens used in the framework of vaccination.

We performed interrupted time series (STI) analysis using the delayed dependent variable model ANCOVA type II Sum Squares with significance for a p-value less than 0.05 to confirm the link between the occurrence of Covid-19 and the collapse of vaccine coverage. These analyzes were performed on global vaccine-preventable disease surveillance data extracted from the District Immunization Data Management Tool (DVD-MT) designed by WHO. Estimates of the target population were obtained from the National Health Information System (SNIS), and surveillance data for Covid-19 patients were obtained from the National Health Security Agency (ANSS).

**Results:** Overall, the EPI recorded a median vaccination coverage of less than 80% for all the vaccines introduced and the analysis of the interrupted time series shows that the interruption of the vaccination program was significant for all the vaccines.

This finding is factual at both the national and district levels. However, there are disparities at this level, even though some districts have yet to report cases of COVID-19 but have experienced drop-in vaccination coverage. The comparison of vaccination coverage for DTP3, for example, shows a sharp drop in the prefectures of Yomou, N’Nzérékoré, Macenta, Kankan, Mandiana, Dinguiraye, Mamou, Koubia, Mali, and Conakry, where it varies between 0 and 80% compared to 2019 where it was above 80%.

**Conclusion:** Our results demonstrate the need for a resilient health system that could adapt quickly and effectively to pandemics and which in turn makes it possible to strengthen EPI activities in Guinea during this period of a health crisis, in particular for children.

## Introduction

Humanity is now facing the COVID-19 pandemic that appeared in China in December 2019. All continents and almost all countries are affected by this “Public Health Emergency of International Concern [1]. This rapidly spreading pandemic has upset the systems, policies, economies, and livelihoods around the world.

In Guinea, the first case was declared on March 12, 2020. Since that date, the number of Covid-19 cases has been increasing day by day despite the state of health emergency and the barrier measures decreed by the Government Guinean. As of June 17, 2020, a total of 4,668 confirmed cases have been registered in the country, including 26 hospital deaths [2].

The effects of COVID-19 pandemic are already starting to be felt economically around the world. According to the recent projections by the International Monetary Fund, global economic losses are expected to be over 10% of global GDP, or more than $ 9 trillion[3]. At the national level in Guinea, in addition to the general slowdown in the production system and the resulting loss of tax revenue, due to the global economic situation and Guinea’s heavy dependence on imports; the country’s GDP is likely to follow the global downward trend due to its reliance on mines and China [4].

In addition to these economic consequences, the global Covid-19 pandemic has not only strained the systems of health in search of resilience but also has affected the continuity of essential services such as immunization programs [5]. It also highlighted the centrality of the work collective of States and WHO to promote health, ensure global security, and serve vulnerable people.

As soon as the WHO announced, drawing on its experience from the Ebola virus disease pandemic, the country developed a COVID-19 operational action plan in February 2020 aimed at preventing and responding if necessary, to this pandemic through the ANSS [6]. This plan was drawn up to coordinate all interventions aimed at strengthening the country’s capacity in response to COVID −19 within the framework of international health regulations. Given the increase in the number of COVID-19 notified cases and the reflection based on the mobility of populations, the socio-political and economic context and the mode of transmission of the disease could lead to the rapid spread of the epidemic [7,8]. The country quickly, the government of the Republic of Guinea, declared a state of health emergency in an address to the nation of the Head of State dated March 27, 2020.

Interruption of routine immunization services could trigger secondary outbreaks of vaccine-preventable diseases and also worsen the long-standing inequity in immunization coverage, especially in rapidly urbanizing cities [9]. For example, due to the COVID-19 pandemic, at least 30 measles vaccination campaigns have been or are at risk of being canceled, which could lead to new outbreaks in 2020 and beyond [10].

Based on these pieces of information, we theorized that the COVID-19 pandemic had a substantial impact on the vaccine activities of the EPI in Guinea and consequently increased the morbidity and mortality of vaccine-preventable diseases. Our study aimed to assess the early impact of COVID-19 on immunization activities by comparing current trends to trends over the past year when immunization coverage improved significantly.

## Methodology

### Place and period of study

The study was carried out in the Extended Immunization Program (EPI) of the Republic of Guinea from February 2020 to June 2020. The EPI is integrated into the pyramidal health system (health post, health center, prefectural hospital, hospital regional, national hospital). The revitalization of the sector materialized with the launch of the first health centers of the Expanded Program of Immunization, Primary Health Care and Essential Medicines (PEV / SSP / ME) from 1988.

### Study design and population

We conducted a retrospective cohort study comparing the trends in administrative coverage of the different antigens used in the context of vaccination through strategies used for the population’s access to vaccines. The periods compared are those before the pandemic (January 2019 to December 2019) and during (from January to June 2020) while taking the month of March 2020, the date on which the pandemic was declared in Guinea as a benchmark.

### Data used

The global vaccine-preventable disease surveillance data used in this study was taken from the District Immunization Data Management Tool (DVD-MT) designed by WHO for data collection and analysis through a configured Excel form. In this file, the data are entered monthly by antigen and by health district and then sent to the EPI through the prefectural and regional health directorates.

Data collection begins at the health post, which is the smallest unit that carries out the vaccination. The monthly data from the health posts are compiled by the health centers, which in turn compile them to send to the DPS level, then the data goes back to the DRS and the EPI. Quality control is done at every step, and feedback is given from the lower level to the central level, where the last compilation is done. The EPI is in the process of migrating to the national surveillance system (DHIS2) set up by the Ministry of Health since 2015. We excluded the data from all the posts for which data was not received at the EPI during the period. Study period.

Estimates of the target population were obtained from the National Health Information System (SNIS), which carried out a general population census in 2014 and which carried out a population projection for the following years until 2025. The Covid-19 surveillance data were obtained from the National Health Security Agency (ANSS), which is responsible for monitoring pandemics in the country.

### Operational definition of variables

The variables of our study are collected at the base by monthly report forms completed by the actors of the districts. The vaccine coverage rates calculated from these variables are based on the indicators present in the EPI performance framework and comply with the new definitions of the concepts used. In our analysis, we used vaccination coverage rates, which are the proportion of people vaccinated in a given population at a given time. Immunization coverage indicators provide an overview of the performance of an immunization system and should be studied in more detail [11]. Although several antigens are administered at the same time of vaccination, we have all taken them into account to see possible rupture caused by COVI-19.The essential antigens, according to the vaccination (table1) schedule, were studied:

**Tableau 1.**
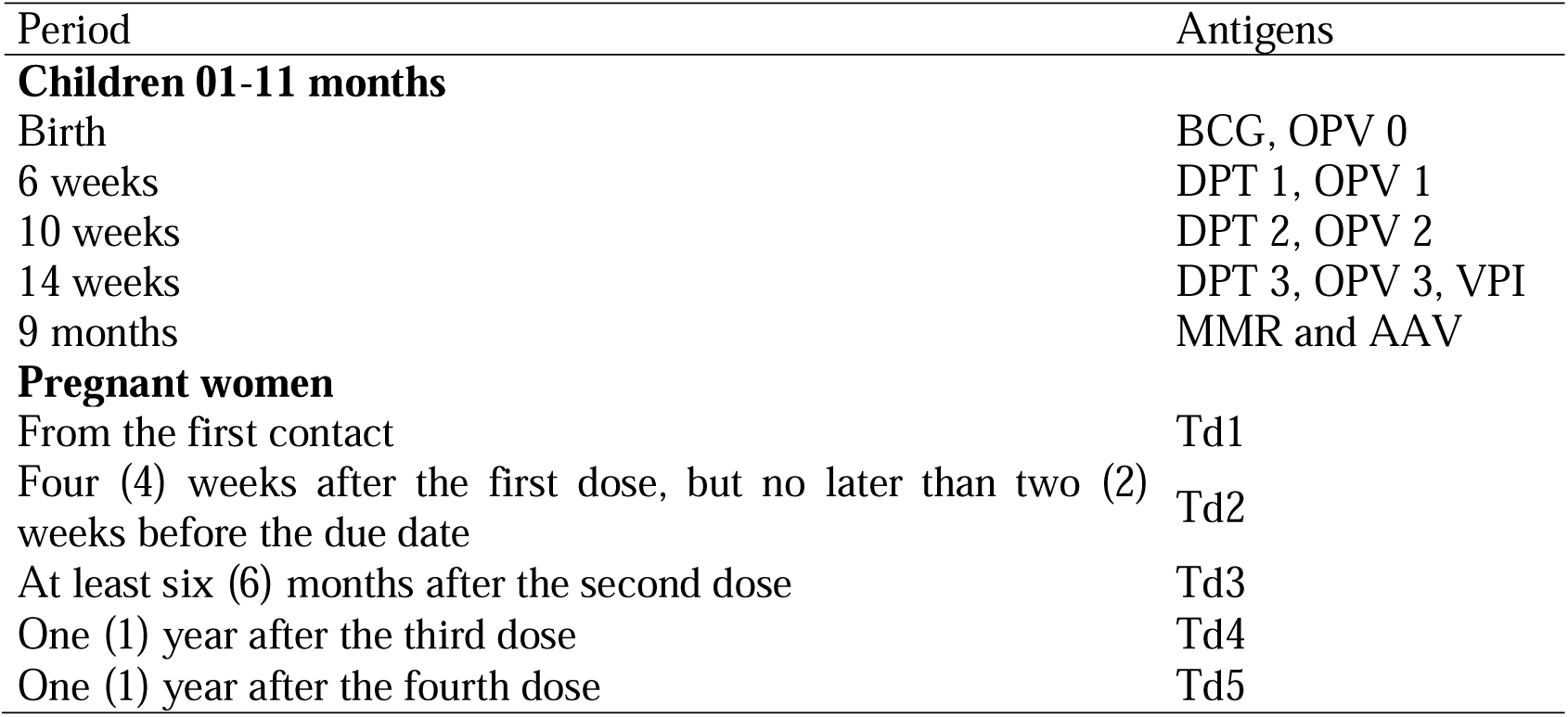
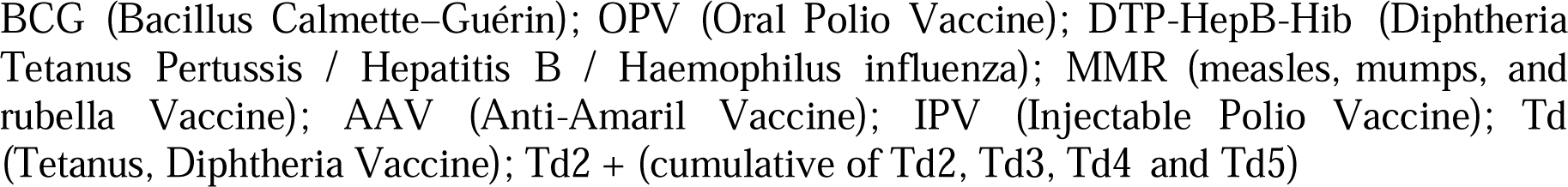
Vaccination schedule for children (0-11 months) and pregnant women

### Statistical analysis plan

A description of the coverage of the main antigens used for vaccination was carried out for the periods 2019 and 2020 through the mean and the standard deviation. A time-series analysis was conducted to assess the effect of Covid-19. To do this, we used the autocorrelation test to examine the significance of the changes observed in each time series, and the cross-correlation to explores the relationship between the COVID-19 series and the vaccine coverage series. Although there are currently meaningless correlations between pairs of independent time series, which are themselves auto-correlated, the link between these two-time series can be explained by the correlation coefficient provided that the two-stationary series [12].

The Dickey-Fuller test was used to verify the stationarity of our time series before their transformation by differentiation (12.13). This differentiation helps remove spurious correlations based on time dependencies between adjacent values in the input time series and removes these influences from the output time series [15]. To confirm and elucidate the observed correlations between time series in the cross-correlation test, we performed an interrupted time series (ITS) analysis using the lagged dependent variable model ANCOVA type II Sum Squares [16]. We have included a default bootstrap model, which runs 1000 replications of the starting model with randomly drawn samples to drive the 95% CI bootstrap. An adjusted F-value (10% suppression) is reported and a p-value is derived. This model was fitted by estimating the average difference of the dependent variables (Vaccine coverage) between the interrupted periods (January to December 2019) and the uninterrupted periods (March to June 2020) by considering the offset of the dependent variable and any other covariates specified in the model [16]. Significance was defined as a p-value of less than 0.05. R 3.5.2 software through the “its.analysis” package was used for the analysis of data compiled in an Excel file.

## Results

Overall over our study period, the EPI recorded a median coverage greater than 70% for all vaccines introduced in Guinea, i.e., 94% for BCG, 92.25% for OPV, 93% for IPV, 95.33 % for DTCs, for MMR, 93% for AAV and 84.5% for Td. We found an average number of fully immunized children (CFI) of 93% (Table 2).

**Table 2.**
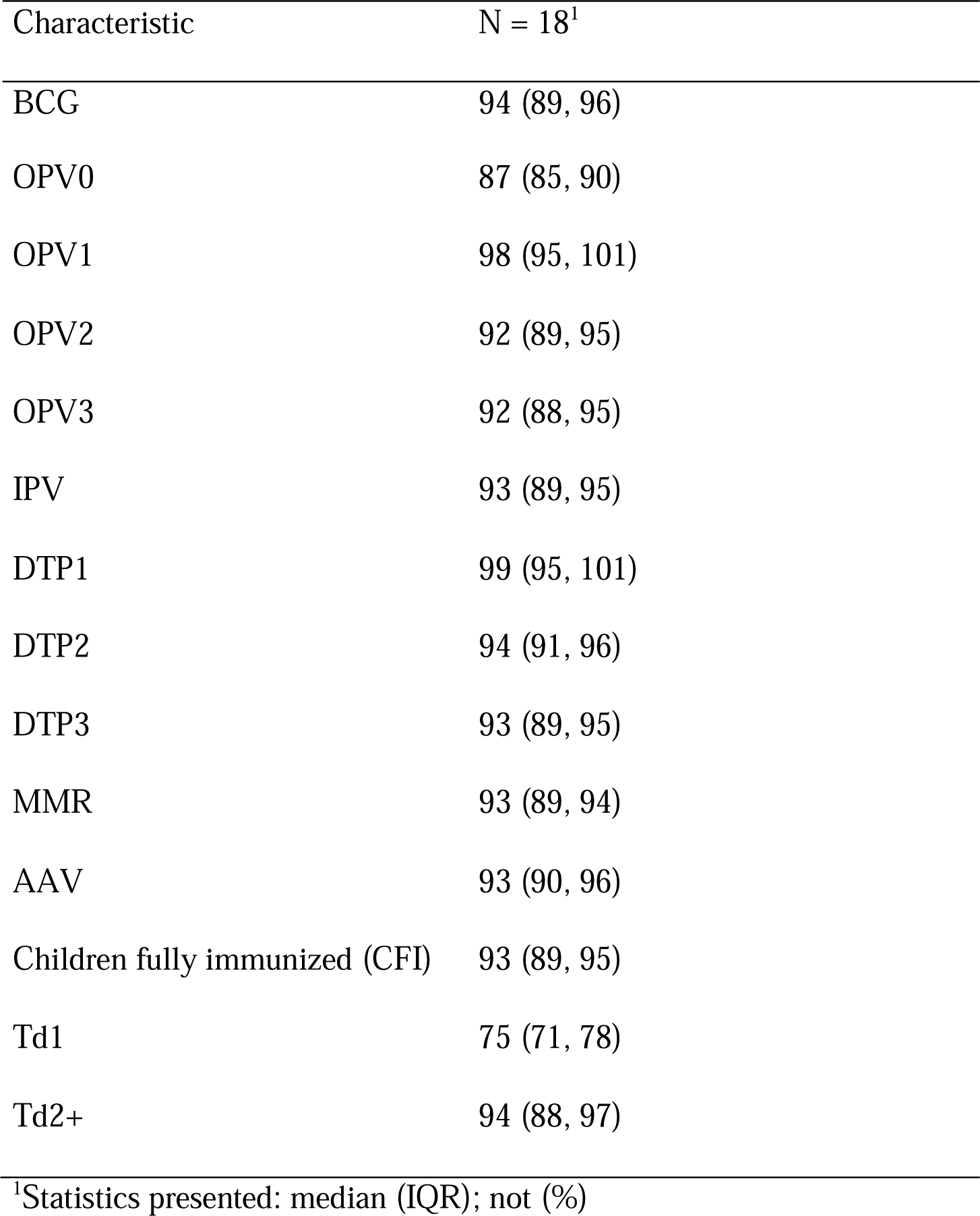
Descriptive statistics of the time series of the main antigens

| Characteristic | N = 18<sup>1</sup> |
| --- | --- |
| BCG | 94 (89, 96) |
| OPV0 | 87 (85, 90) |
| OPV1 | 98 (95, 101) |
| OPV2 | 92 (89, 95) |
| OPV3 | 92 (88, 95) |
| IPV | 93 (89, 95) |
| DTP1 | 99 (95, 101) |
| DTP2 | 94 (91, 96) |
| DTP3 | 93 (89, 95) |
| MMR | 93 (89, 94) |
| AAV | 93 (90, 96) |
| Children fully immunized (CFI) | 93 (89, 95) |
| Td1 | 75 (71, 78) |
| Td2+ | 94 (88, 97) |

<sup>1</sup>Statistics presented: median (IQR); not (%)

A comparative analysis of fourteen months (14) months before the declaration of the pandemic in Guinea and during the first four months of the pandemic shows a downward trend in vaccination coverage for all antigens. This decrease is much more marked for IPV and DTP3, i.e., 94 before the pandemic and 73 during the pandemic (Table 3 and Figure 1,2,3). A similar trend was observed for the number of children fully immunized among and pregnant women (Table 2 and Figures 2 and 3). This result can be seen in the graphs of the cross-correlation test, which illustrate significant negative shifts for most antigens (figure 4, figure 5, and figure 5) except OPV 0 (figure 4, A) and TD2 + (figure 5, K).

**Table 3.** Median vaccination coverage of the main antigens before and during the Covid-19 pandemic

| Characteristic | Before, N = 14<sup>1</sup> | during, N = 4<sup>1</sup> |
| --- | --- | --- |
| BCG | 95 (91, 97) | 77 (71, 86) |
| OPV0 | 88 (86, 90) | 72 (66, 81) |
| OPV1 | 99 (95, 102) | 79 (71, 89) |
| OPV2 | 94 (90, 96) | 74 (67, 84) |
| OPV3 | 93 (89, 96) | 73 (66, 83) |
| IPV | 94 (90, 96) | 73 (66, 83) |
| DTP1 | 99 (98, 102) | 79 (72, 89) |
| DTP2 | 94 (92, 97) | 75 (67, 84) |
| DTP3 | 94 (91, 96) | 73 (67, 83) |
| MMR | 93 (92, 95) | 74 (67, 83) |
| AAV | 94 (91, 97) | 74 (67, 83) |
| Children fully immunized (CFI) | 94 (91, 97) | 73 (66, 82) |
| Td1 | 77 (74, 78) | 62 (59, 65) |
| Td2+ | 95 (92, 98) | 75 (69, 84) |

<sup>1</sup>Statistics presented: median (IQR); n (%)

**Figure 1.**
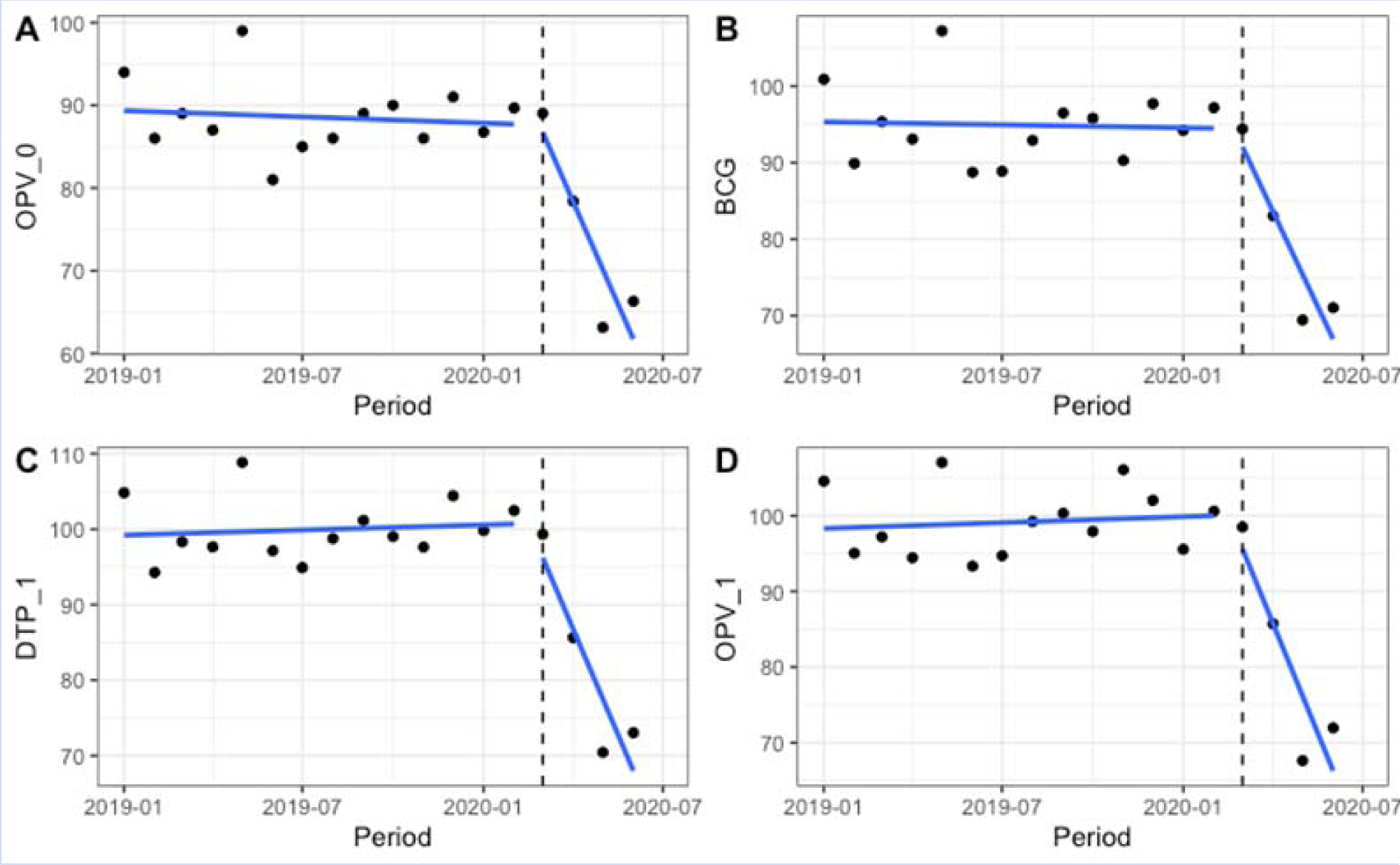
Trends in vaccine coverage for (a) OPV_0, (b) BCG, (c) DTP1, and (d) OPV1 antigens

**Figure 2.**
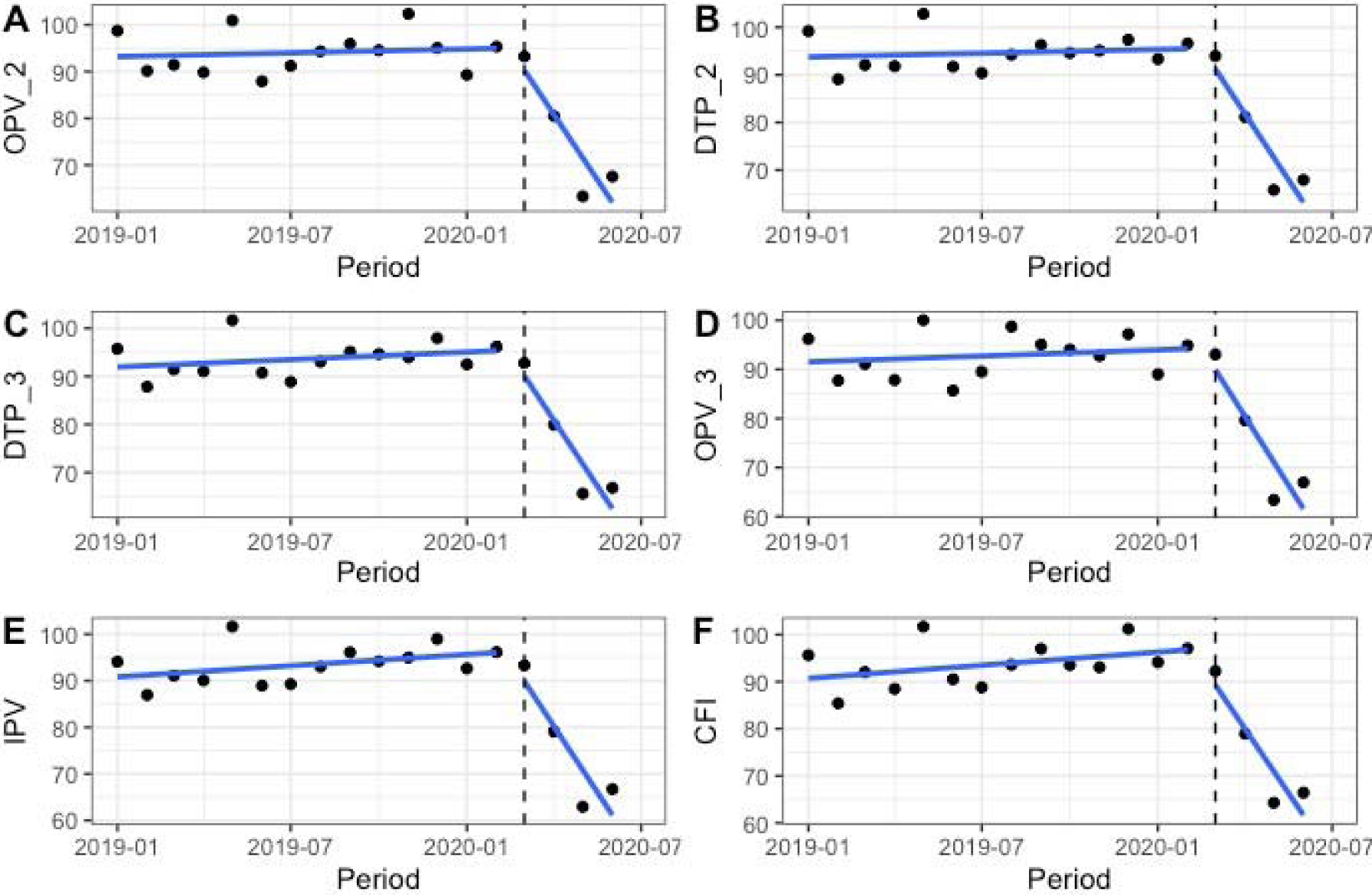
Trends in vaccine coverage of antigens (a) OPV_2, (b) DTP2, (c) DTP3, and (d) OPV3, (e) IPV, (f) Children fully vaccinated (CFI)

**Figure 3.**
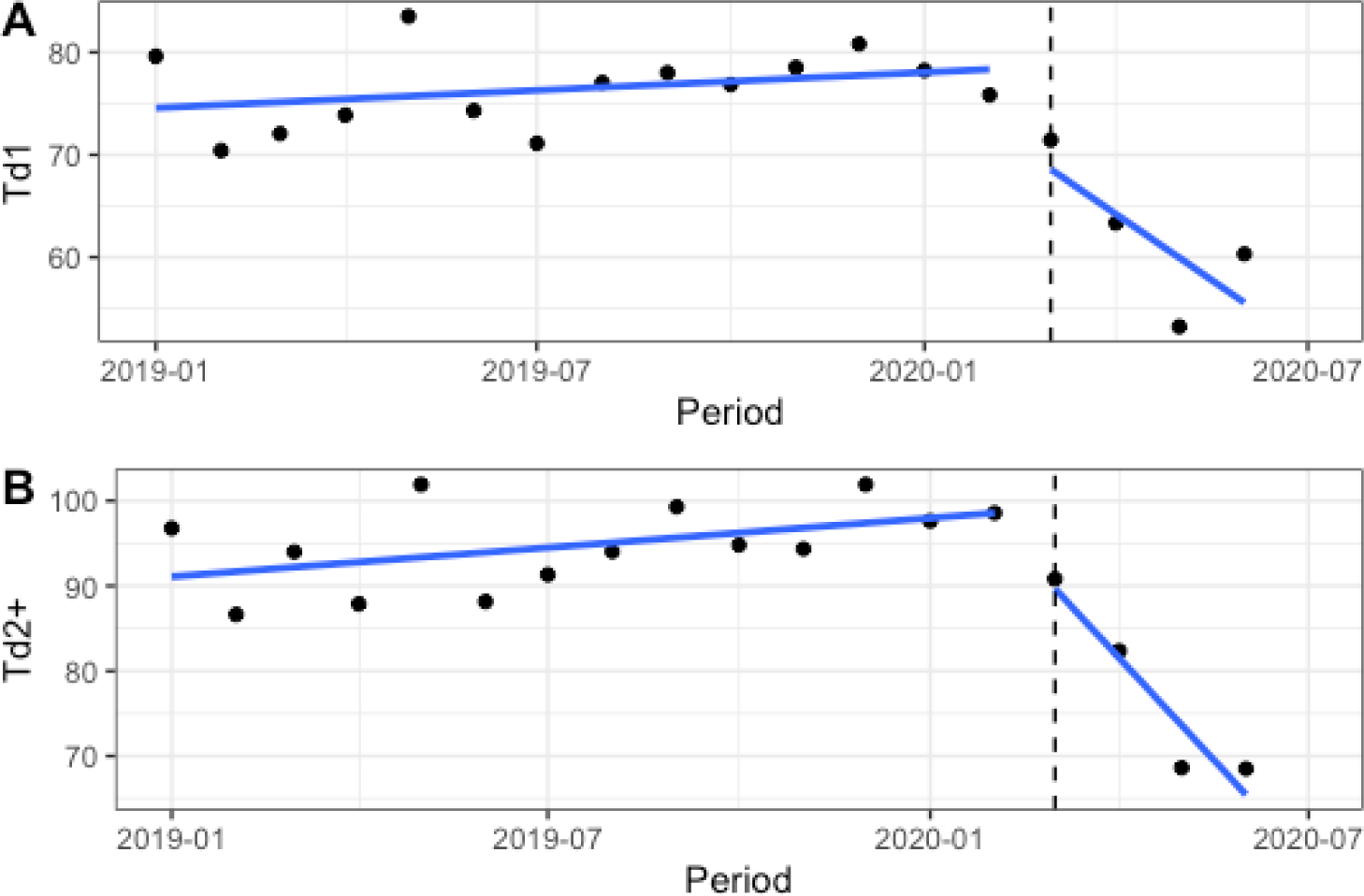
Trends in vaccine coverage of antigens (a) Td1 and (b) Td2

**Figure 4.**
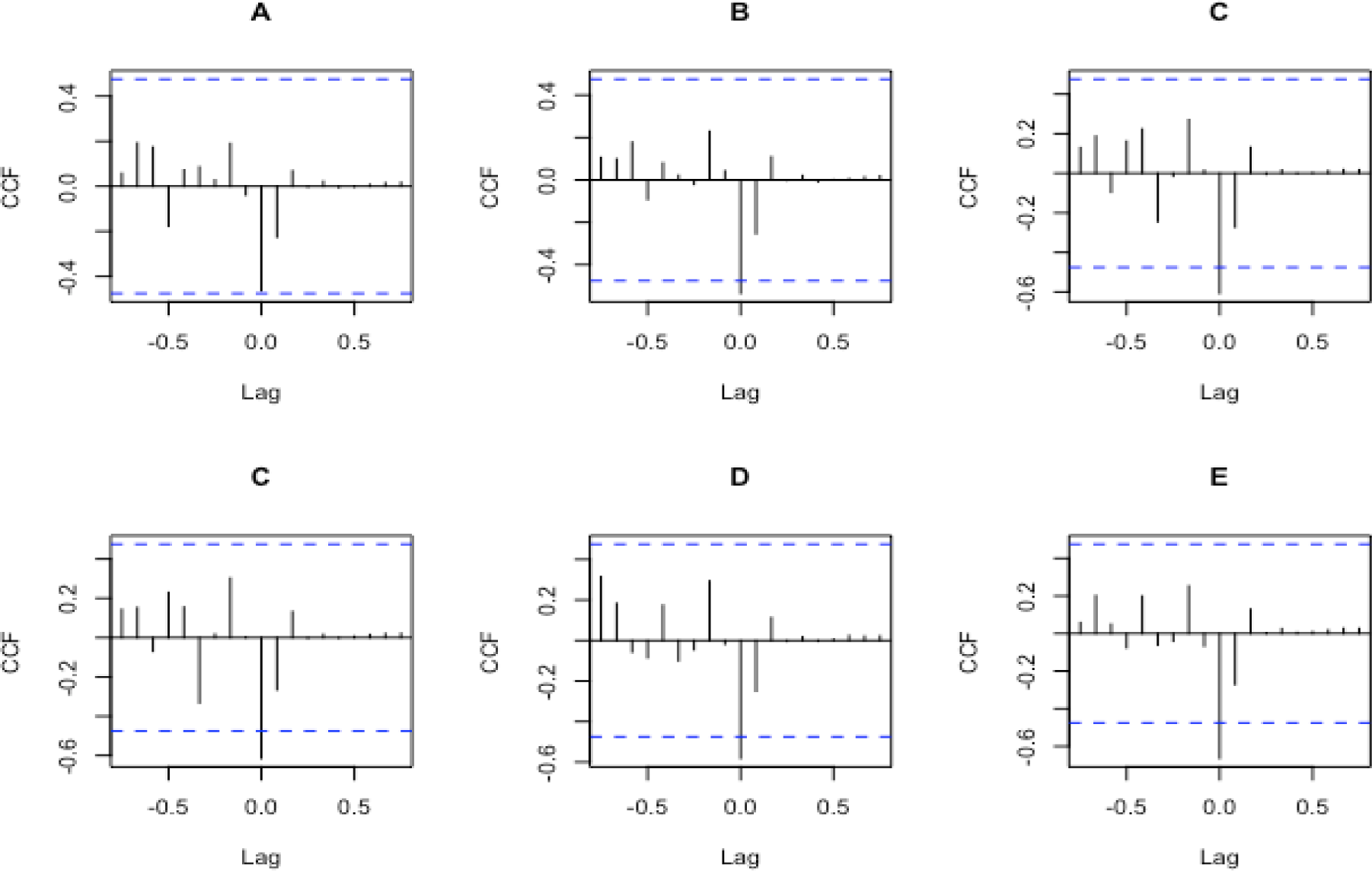
Cross-correlation test for COVID-19 time series and antigens (a) OPV_0, (b) BCG, (c) DTP1, (d) OPV_1 (e) and OPV_2

**Figure 5.**
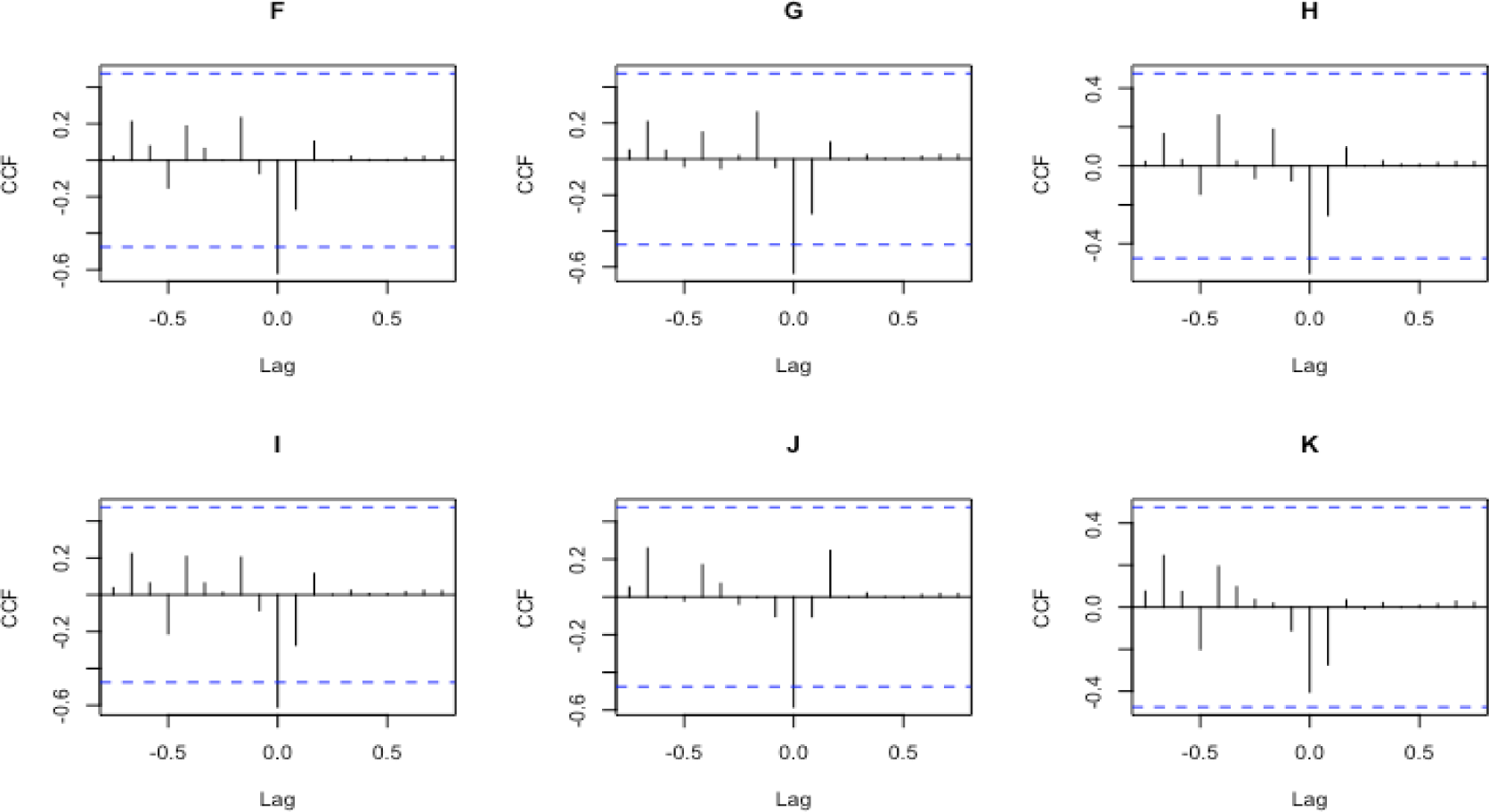
Cross-correlation test for COVID-19 time series and antigens (f) OPV_2, (g) DTP2, (h)OPV3, (i) IPV, (j) Td1, (k) Td2+

**Figure 6.**
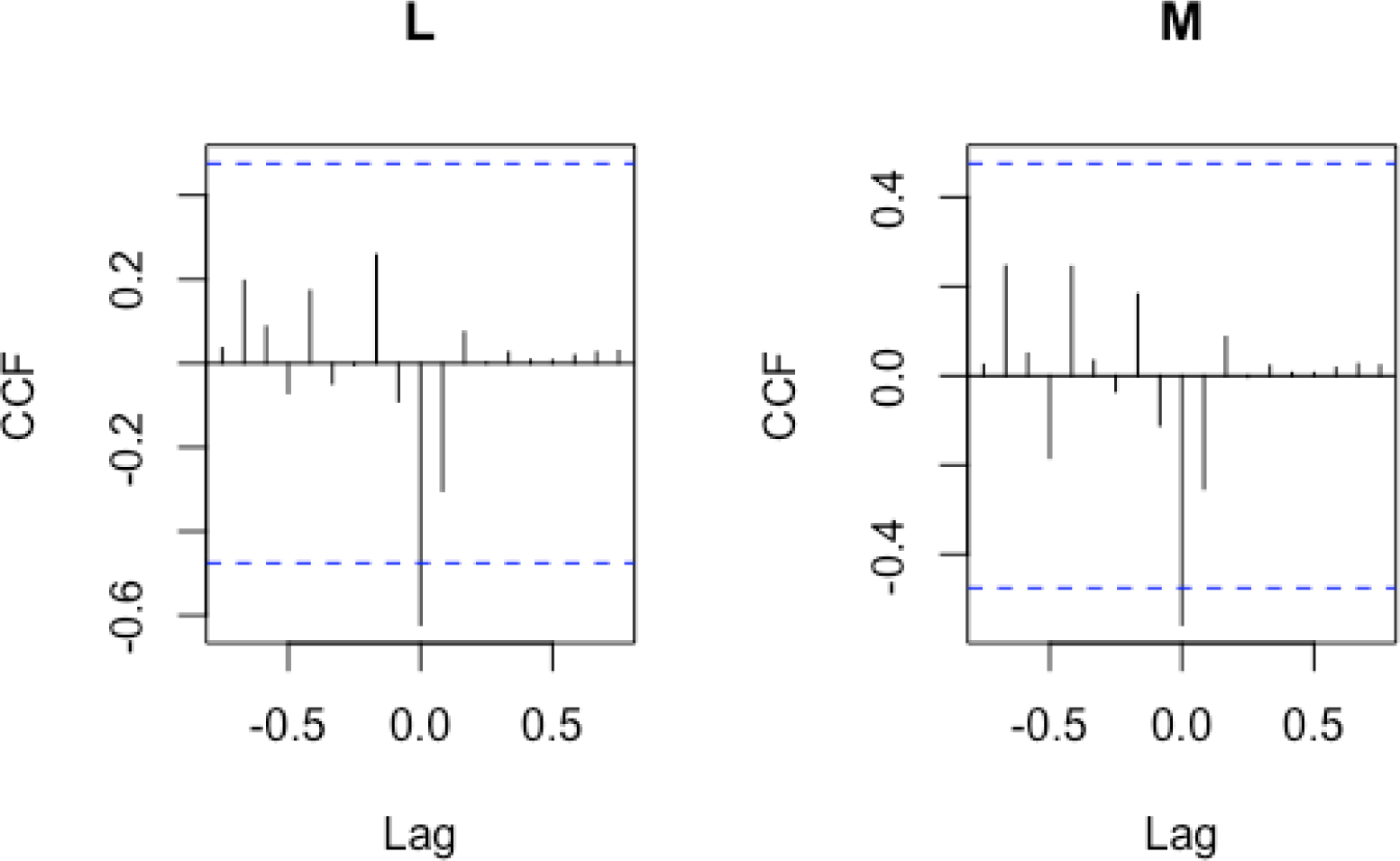
Cross-correlation test for COVID-19 time series and antigens (m) DTP3 and (l) Children fully vaccinated

The analysis of interrupted time series shows that the interruption of the vaccination program was significant for all the vaccines, however this interruption resulted in a significant delay for four vaccines: IPV (F value = 23.69 [95% CI 3.07 to 57.69], P = 0.0002), DTP1 (F value = 21.77 [95% CI 3.36 to 78.39], P = 0.00036), DTP2 (F value = 25.30 [95% CI 3.56 to 97.49], P = 0.0002), DTP3 (F value = 24.49 [95% CI 4.27 to 61.92], P = 0.0002) and AAV (F value = 19.01 [95% CI 3.89 to 69.35], P = 0.00065) (Table 4).

**Table 4.**
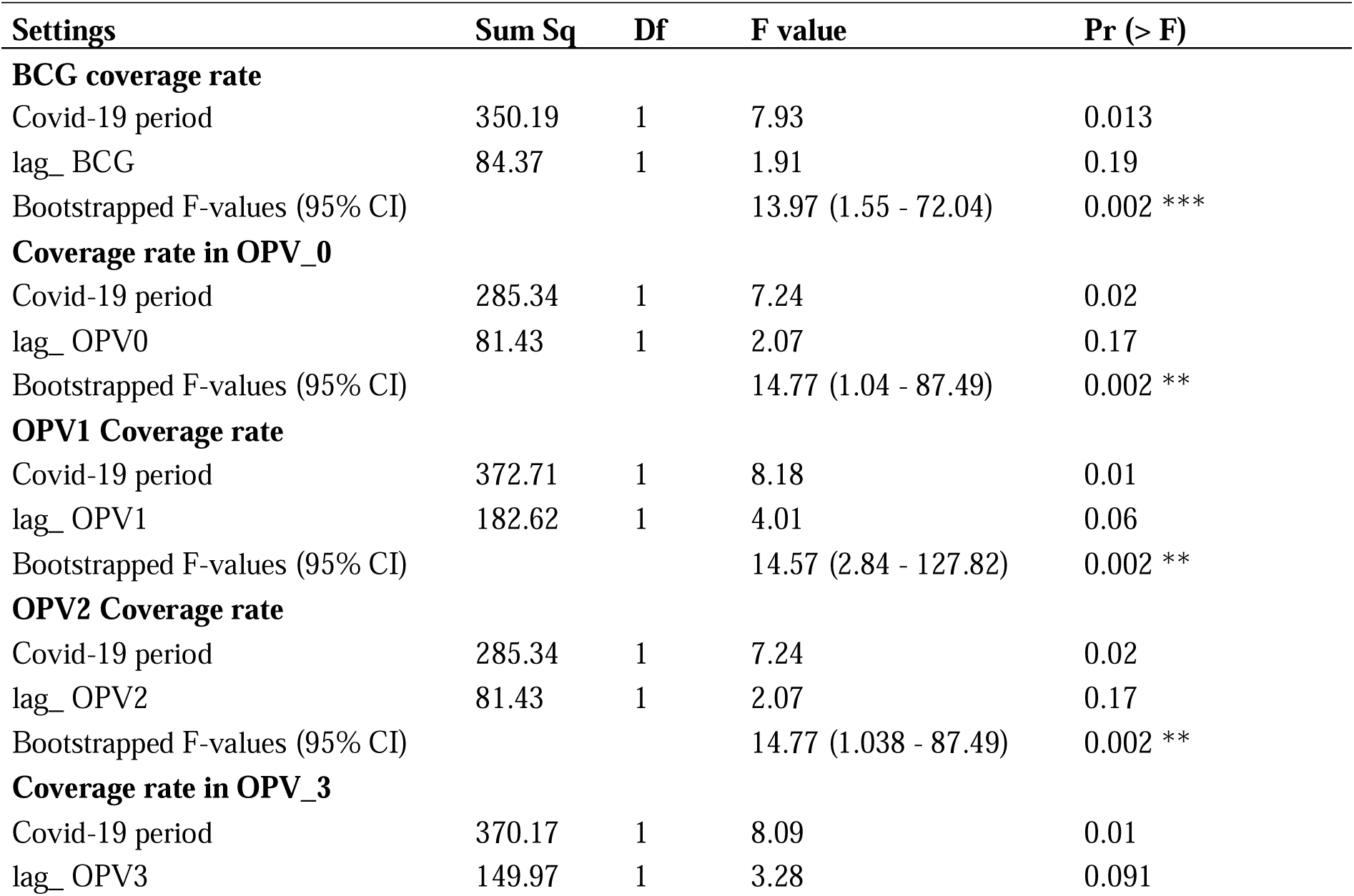
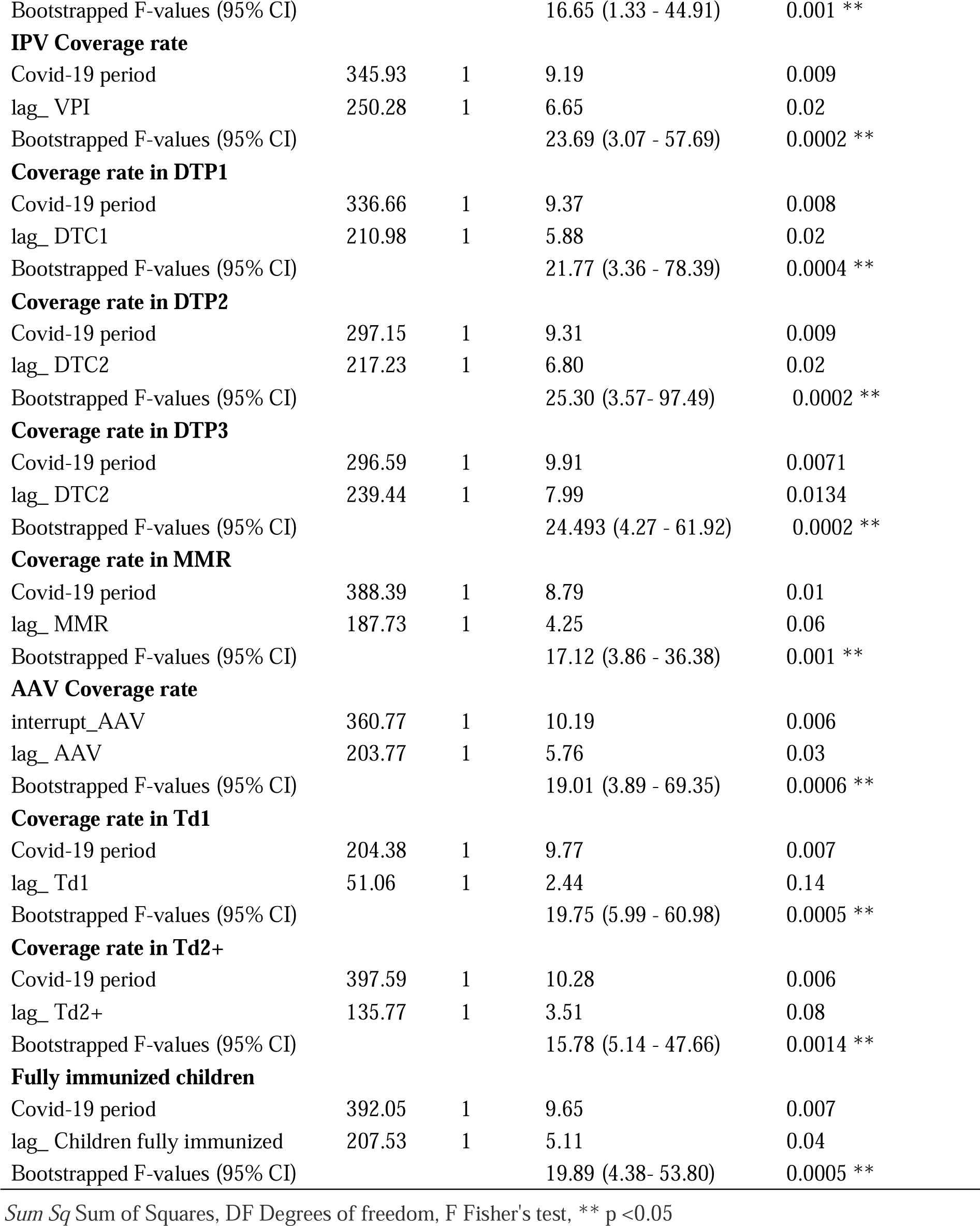
Results of the analysis model of the interrupted time series of vaccine coverage during the Covid-19 pandemic

| Settings | Sum Sq | Df | F value | Pr (> F) |
| --- | --- | --- | --- | --- |
| **BCG coverage rate** |  |  |  |  |
| Covid-19 period | 350.19 | 1 | 7.93 | 0.013 |
| lag_ BCG | 84.37 | 1 | 1.91 | 0.19 |
| Bootstrapped F-values (95% CI) |  |  | 13.97 (1.55 - 72.04) | 0.002 *** |
| **Coverage rate in OPV_0** |  |  |  |  |
| Covid-19 period | 285.34 | 1 | 7.24 | 0.02 |
| lag_ OPV0 | 81.43 | 1 | 2.07 | 0.17 |
| Bootstrapped F-values (95% CI) |  |  | 14.77 (1.04 - 87.49) | 0.002 ** |
| **OPV1 Coverage rate** |  |  |  |  |
| Covid-19 period | 372.71 | 1 | 8.18 | 0.01 |
| lag_ OPV1 | 182.62 | 1 | 4.01 | 0.06 |
| Bootstrapped F-values (95% CI) |  |  | 14.57 (2.84 - 127.82) | 0.002 ** |
| **OPV2 Coverage rate** |  |  |  |  |
| Covid-19 period | 285.34 | 1 | 7.24 | 0.02 |
| lag_ OPV2 | 81.43 | 1 | 2.07 | 0.17 |
| Bootstrapped F-values (95% CI) |  |  | 14.77 (1.038 - 87.49) | 0.002 ** |
| **Coverage rate in OPV_3** |  |  |  |  |
| Covid-19 period | 370.17 | 1 | 8.09 | 0.01 |
| lag_ OPV3 | 149.97 | 1 | 3.28 | 0.091 |
| Bootstrapped F-values (95% CI) |  |  | 16.65 (1.33 - 44.91) | 0.001 ** |
| **IPV Coverage rate** |  |  |  |  |
| Covid-19 period | 345.93 | 1 | 9.19 | 0.009 |
| lag_ VPI | 250.28 | 1 | 6.65 | 0.02 |
| Bootstrapped F-values (95% CI) |  |  | 23.69 (3.07 - 57.69) | 0.0002 ** |
| **Coverage rate in DTP1** |  |  |  |  |
| Covid-19 period | 336.66 | 1 | 9.37 | 0.008 |
| lag_ DTC1 | 210.98 | 1 | 5.88 | 0.02 |
| Bootstrapped F-values (95% CI) |  |  | 21.77 (3.36 - 78.39) | 0.0004 ** |
| **Coverage rate in DTP2** |  |  |  |  |
| Covid-19 period | 297.15 | 1 | 9.31 | 0.009 |
| lag_ DTC2 | 217.23 | 1 | 6.80 | 0.02 |
| Bootstrapped F-values (95% CI) |  |  | 25.30 (3.57- 97.49) | 0.0002 ** |
| **Coverage rate in DTP3** |  |  |  |  |
| Covid-19 period | 296.59 | 1 | 9.91 | 0.0071 |
| lag_ DTC2 | 239.44 | 1 | 7.99 | 0.0134 |
| Bootstrapped F-values (95% CI) |  |  | 24.493 (4.27 - 61.92) | 0.0002 ** |
| **Coverage rate in MMR** |  |  |  |  |
| Covid-19 period | 388.39 | 1 | 8.79 | 0.01 |
| lag_ MMR | 187.73 | 1 | 4.25 | 0.06 |
| Bootstrapped F-values (95% CI) |  |  | 17.12 (3.86 - 36.38) | 0.001 ** |
| **AAV Coverage rate** |  |  |  |  |
| interrupt_AAV | 360.77 | 1 | 10.19 | 0.006 |
| lag_ AAV | 203.77 | 1 | 5.76 | 0.03 |
| Bootstrapped F-values (95% CI) |  |  | 19.01 (3.89 - 69.35) | 0.0006 ** |
| **Coverage rate in Td1** |  |  |  |  |
| Covid-19 period | 204.38 | 1 | 9.77 | 0.007 |
| lag_ Td1 | 51.06 | 1 | 2.44 | 0.14 |
| Bootstrapped F-values (95% CI) |  |  | 19.75 (5.99 - 60.98) | 0.0005 ** |
| **Coverage rate in Td2+** |  |  |  |  |
| Covid-19 period | 397.59 | 1 | 10.28 | 0.006 |
| lag_ Td2+ | 135.77 | 1 | 3.51 | 0.08 |
| Bootstrapped F-values (95% CI) |  |  | 15.78 (5.14 - 47.66) | 0.0014 ** |
| **Fully immunized children** |  |  |  |  |
| Covid-19 period | 392.05 | 1 | 9.65 | 0.007 |
| lag_ Children fully immunized | 207.53 | 1 | 5.11 | 0.04 |
| Bootstrapped F-values (95% CI) |  |  | 19.89 (4.38- 53.80) | 0.0005 ** |

*Sum Sq* Sum of Squares, *DF* Degrees of freedom, *F* Fisher's test, \*\* $p < 0.05$

At the district level, the comparison of half-yearly vaccination coverage for DTP3 for the years 2019 and 2020 (map 1 and 2), for example, shows a sharp drop in the prefectures of Yomou, N’Zérékoré, Macenta, Kankan, Mandiana, Dinguiraye, Mamou, Koubia, Mali and Conakry, where it varied between 0 and 80% compared to 2019 when it was above 80%. On the other hand, the prefecture of Forécariah saw an improvement in 2020, where the coverage in DTP3 is higher than 80%.

## Discussions

This comparative analysis between the months preceding the pandemic in Guinea and the first months of the pandemic shows a downward trend in vaccination coverage of all antigens. According to data collected by the World Health Organization, UNICEF, Gavi, and the Sabin Vaccine Institute, the delivery of routine immunization services is significantly hampered in at least 68 countries and will likely affect around 80 million fewer children 1-year old living in these countries. Since March 2020, routine childhood immunization services have been disrupted globally [10]. Authors have also reported a decline in the trend in vaccine coverage during the COVID-19 pandemic, particularly in the USA [17], in England [18], and Israel [19]. African countries are not spared in this observation; According to a new survey by UNICEF, WHO, and Gavi, conducted in collaboration with the US Centers for Disease Control, the Sabin Vaccine Institute, and the Johns Hopkins Bloomberg School of Public Health, the three-quarters of the 82 countries that participated reported COVID-19-related disruptions in their immunization programs in May 2020 [20].

Our results show that the decrease is much more marked for vaccines, IPV, and DTP3. This result is similar to that observed in England, where the drop-in coverage of the hexavalent vaccine is more significant in 2020 (the change in coverage was −3.5 (95% CI −3.7 to −3.4) during the pandemic of COVID-19. The change was also harmful to the MMR, where the change was −3.7 (95% CI −3.8 to −3.6)[18]. In contrast, data from the US Vaccine Tracking System indicates a notable decrease in orders for childhood influenza vaccines and measles vaccines compared to 2019 [21]. The fear caused by the number of deaths by COVID-19 in China and Europe, the restriction of travel, the increase in transport costs, the compulsory wearing of a face mask, the distancing measures imposed by the Guinean authorities associated with Insufficient communication channels have led to a reduction in the number of people going to health services, including vaccination.

Our results show that the interruption of the vaccine program was significant for all vaccines. However, this interruption resulted in a significant delay in the program than for IPV, DTP1, DTP2, DTP3, and AAV. Most field activities (training, supervision, routine vaccination, and advanced strategy, etc.) were temporarily suspended. It must be recognized that attendance has fallen sharply, which can be observed through all the program indicators. Some parents are still reluctant to leave the house due to movement restrictions, lack of information, or because they fear they may be infected with the COVID-19 virus. And many health workers are unavailable due to restrictions on travel or redeployment to COVID response functions [10].

Based on the conclusions of this study Abbass et al. [22], deaths averted by supporting routine childhood immunizations in Africa outweigh the excessive risk of death from COVID-19 associated with visits to immunization clinics, especially for vaccinated children. All suitable measures should be put in place to promote vaccination, even if they would alleviate certain constraints related to the state of health emergency in the COVID-19 context.

## Conclusion

This study shows a significant negative relationship between the vaccination coverage of the main antigens and the occurrence of COVI-19 despite the measures taken by the Guinean State to guarantee the continuity of services and the awareness of the population to its adherence to vaccination in the context of COVID-19. Our results persuade the Guinean government through the Ministry of Health to effectively improve the response system to the pandemic, although efforts have already been made regarding the Ebola virus disease. The establishment of a specific and compelling emergency mechanism to support large-scale programs such as the Expanded Program on Immunization may also be necessary.

## Supporting information

R script

## Data Availability

Data are available on request from the Expanded Program on Immunization (EPI) of Guinea. The statistical analysis carried out in our study is fully available and can be reproduced if necessary, through the R code attached to this article.

## Statements

### Ethical approval and consent to participate

This study used aggregate immunization surveillance data collected from registers at health centers across the country. Authorization from the Guinean Ministry of Health through the EPI was obtained for data analysis within the strict framework of an evaluation of immunization services.

### Consent to publication

Not applicable

### Competing interests

The authors declare that they have no competing interest in this work.

### Funding

This study did not receive any specific funding apart from technical assistance provided by WHO Guinea, which is a strategic partner of the EPI.

### Contributions from authors

DM emitted the idea of the study, contributed to the study design, participated in the analysis and interpretation of the data and wrote the manuscript, SY and MK contributed to the design, organization of the research, manuscript writing and critical review, DS, AC, BM, and SS, contributed to the study design, organization of the research project, supervision of data collection and critical review of the manuscript, IB, LF, YS, IC, GC and AT commented on the manuscript. AM and AT contributed to the methodology and performed the data analysis and corrected the manuscript. All authors have approved the final version of the manuscript.

## Acknowledgements

We thank the Expanded Program on Immunization (EPI) for their collaboration and the GAVI Alliance and WHO for their daily support in the eradication of vaccine-preventable diseases.

---

**Map1.**
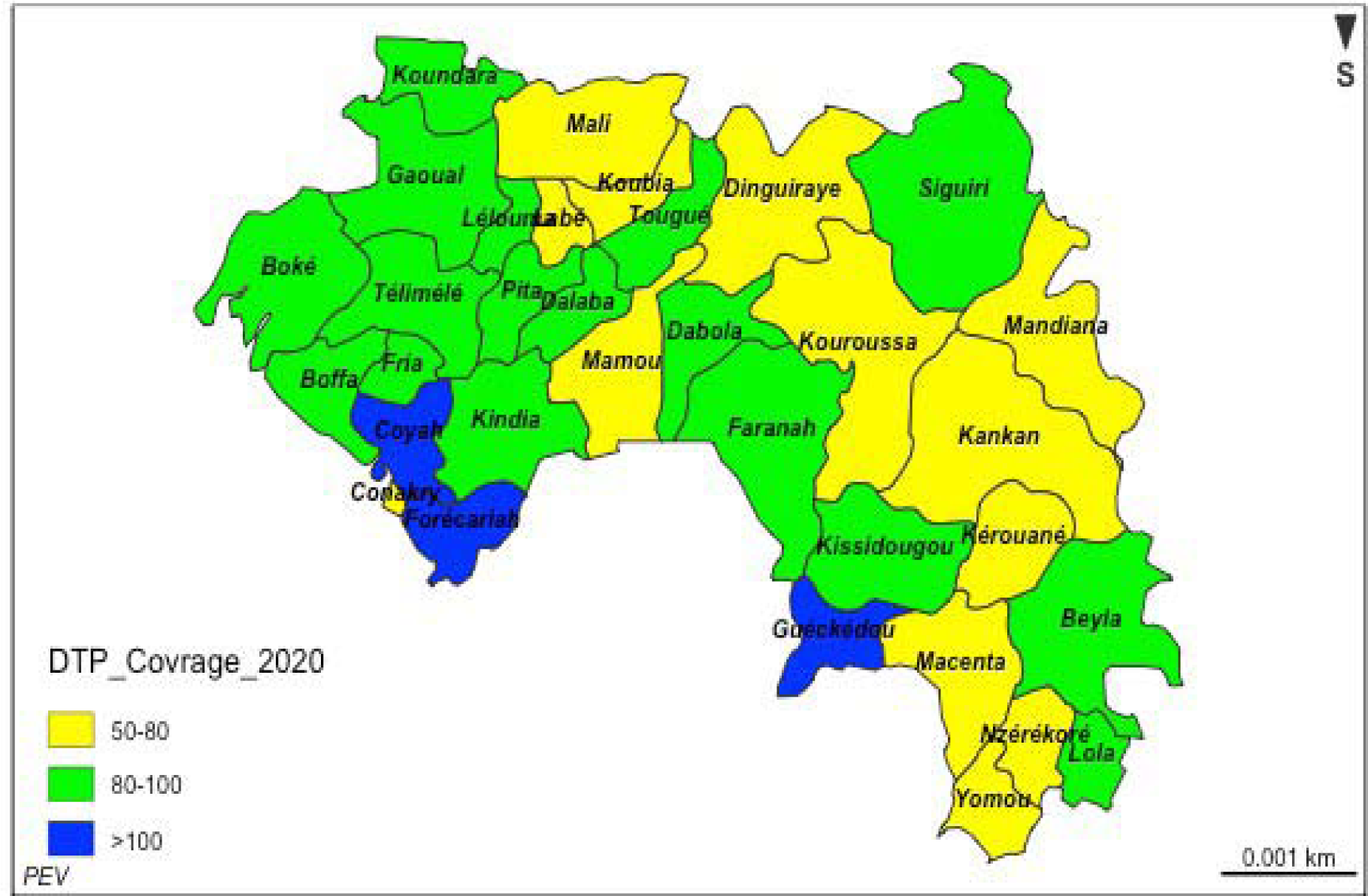
half-yearly coverage for DTP 3 antigen in 2020

**Map1.**
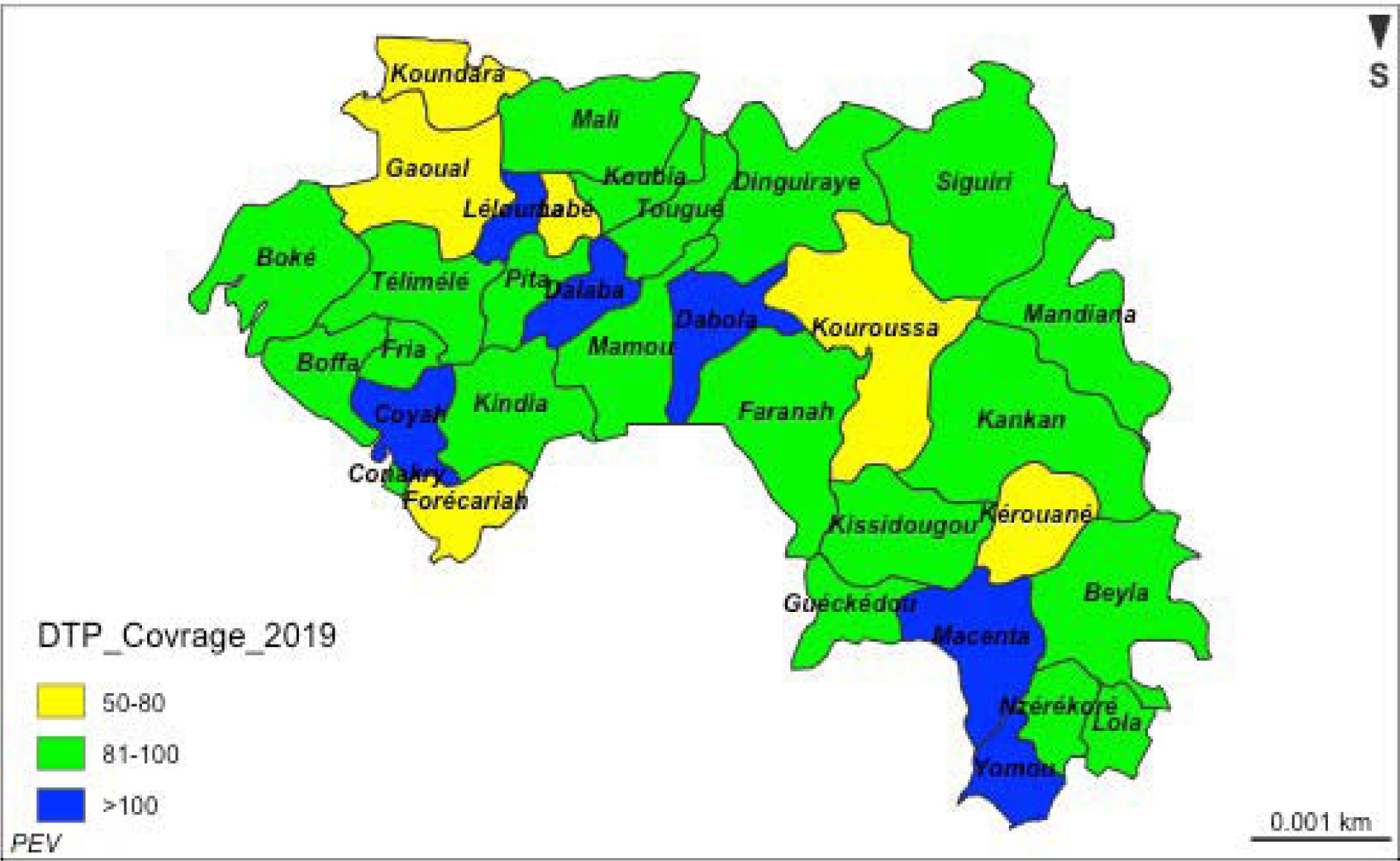
half-yearly coverage for DTP 3 antigen in 2019

## Notes

### Competing Interest Statement

The authors have declared no competing interest.

### Funding Statement

This study did not receive any specific funding apart from technical assistance provided by Guinea, which is a strategic partner of the EPI.

### Author Declarations

Ministry of Health (Service evaluation)

## References

1. WHO. COVID-19 as a Public Health Emergency of International Concern (PHEIC) under the IHR | Strategic Partnership for IHR and Health Security (SPH) [Internet]. 2020 [cité 2020 août 20]; Available from: https://extranet.who.int/sph/covid-19-public-health-emergency-international-concern-pheic-under-ihr

2. ANSS. Continuité des services de santé en période de COVID-19 en Guinée [Internet]. 2020 [cité 2020 août 20];Available from: https://anss-guinee.org/Article/singlePost/726

3. International Monetary Fund. World Economic Outlook, April 2020: The Great Lockdown [Internet]. IMF2020 [cité 2020 août 20];Available from: https://www.imf.org/en/Publications/WEO/Issues/2020/04/14/weo-april-2020

4. Gouverment Guinéen. Plan de riposte économique à la crise sanitaire COVID-19. 2020;

5. Dzinamarira T, Dzobo M, Chitungo I. COVID-19: A perspective on Africa’s capacity and response. J. Med. Virol. [Internet] 2020 [cité 2020 août 20];n/a. Available from: https://onlinelibrary.wiley.com/doi/abs/10.1002/jmv.26159

6. Maxmen A. Ebola prepared these countries for coronavirus — but now even they are floundering. Nature [Internet] 2020 [cité 2020 août 20];583:667□8. Available from: https://www.nature.com/articles/d41586-020-02173-z

7. Chen Z-L, Zhang Q, Lu Y, Guo Z-M, Zhang X, Zhang W-J, et al. Distribution of the COVID-19 epidemic and correlation with population emigration from Wuhan, China. Chin. Med. J. (Engl.) 2020; 133:1044□50.

8. Jiang J, Luo L. Influence of population mobility on the novel coronavirus disease (COVID-19) epidemic: based on panel data from Hubei, China. Glob. Health Res. Policy [Internet] 2020 [cité 2020 août 20];5. Available from: https://www.ncbi.nlm.nih.gov/pmc/articles/PMC7276249/

9. Chandir S, Siddiqi DA, Setayesh H, Khan AJ. Impact of COVID-19 lockdown on routine immunisation in Karachi, Pakistan. Lancet Glob. Health [Internet] 2020 [cité 2020 août 20];8: e1118L20. Available from: https://www.ncbi.nlm.nih.gov/pmc/articles/PMC7324087/

10. WHO. At least 80 million children under one at risk of diseases such as diphtheria, measles and polio as COVID-19 disrupts routine vaccination efforts, warn Gavi, WHO and UNICEF [Internet]. 2020. Available from: https://www.who.int/news-room/detail/22-05-2020-at-least-80-million-children-under-one-at-risk-of-diseases-such-as-diphtheria-measles-and-polio-as-covid-19-disrupts-routine-vaccination-efforts-warn-gavi-who-and-unicef

11. WHO. Guide pour l’harmonisation des indicateurs de couverture vaccinale dans le cadre des enquêtes auprès des ménages. 2020;

12. Probst WN, Stelzenmüller V, Fock HO. Using cross-correlations to assess the relationship between time-lagged pressure and state indicators: an exemplary analysis of North Sea fish population indicators. ICES J. Mar. Sci. [Internet] 2012 [cité 2020 août 31];69:670□81. Available from: https://academic.oup.com/icesjms/article/69/4/670/632477

13. Dean RT, Dunsmuir WTM. Dangers and uses of cross-correlation in analyzing time series in perception, performance, movement, and neuroscience: The importance of constructing transfer function, autoregressive models. Behav. Res. Methods [Internet] 2016 [cité 2020 août 31]; 48:783□802. Available from: https://doi.org/10.3758/s13428-015-0611-2

14. Zeger SL, Irizarry R, Peng RD. ON-TIME SERIES ANALYSIS OF PUBLIC HEALTH AND BIOMEDICAL DATA. Annu. Rev. Public Health [Internet] 2006 [cité 2020 août 31]; 27:57□79. Available from: http://www.annualreviews.org/doi/10.1146/annurev.publhealth.26.021304.144517

15. Patel AX, Kundu P, Rubinov M, Jones PS, Vértes PE, Ersche KD, et al. A wavelet method for modeling and despising motion artifacts from resting-state fMRI time series. NeuroImage [Internet] 2014 [cité 2020 août 31];95:287□304. Available from: http://www.sciencedirect.com/science/article/pii/S1053811914001578

16. English P. The its.analysis R Package – Modelling Short Time Series Data [Internet]. Rochester, NY: Social Science Research Network; 2019 [cité 2020 août 31]. Available from: https://papers.ssrn.com/abstract=3398189

17. Szablewski CM. SARS-CoV-2 Transmission and Infection Among Attendees of an Overnight Camp [Internet]. Georgia: 2020. Available from: https://www.cdc.gov/mmwr/volumes/69/wr/mm6931e1.htm

18. McDonald HI, Tessier E, White JM, Woodruff M, Knowles C, Bates C. Early impact of the coronavirus disease (COVID-19) pandemic and physical distancing measures on routine childhood vaccinations in England. Eurosurveillance 2020;14 mai 2020;25(19):2000848.

19. Personnel du Times of Israel. Vaccinations drop amid COVID-19 fears, raising specter of fresh measles outbreak [Internet]. TIMES OF ISRAEL; Available from: https://www.timesofisrael.com/vaccinations-drop-amid-virus-fears-raising-specter-of-fresh-measles-outbreak/

20. WHO. L’OMS et l’UNICEF mettent en garde contre une baisse de la vaccination pendant la pandémie de COVID-19 [Internet]. 2020 [cité 2020 sept 5];Available from: https://www.who.int/fr/news-room/detail/15-07-2020-who-and-unicef-warn-of-a-decline-in-vaccinations-during-covid-19

21. Santoli JM, Lindley MC, DeSilva MB, Kharbanda EO, Daley MF, Galloway L. Effects of the COVID-19 Pandemic on Routine Pediatric Vaccine Ordering and Administration — United States, 2020. MMWR Morb Mortal Wkly Rep 2020; 69:591–3.

22. Abbas K, Procter SR, Zandvoort K van, Clark A, Funk S, Mengistu T, et al. Routine childhood immunisation during the COVID-19 pandemic in Africa: a benefit-risk analysis of health benefits versus excess risk of SARS-CoV-2 infection. Lancet Glob. Health [Internet] 2020 [cité 2020 sept 5];0. Available from: https://www.thelancet.com/journals/langlo/article/PIIS2214-109X(20)30308-9/abstract

