## Supplementary material for "Early effect of the Covid-19 epidemic on vaccine coverage of major antigens in Guinea: an analysis of the interrupted time series of national immunization coverage": R script: script.R

```
library(readxl)
BDD_PEV <- read_excel("~/Desktop/BDD_PEV.xlsx", 
                      col_types = c("date","numeric", "numeric", "numeric", 
                                    "numeric", "numeric", "numeric", 
                                    "numeric", "numeric", "numeric", 
                                    "numeric", "numeric", "numeric", 
                                    "numeric", "numeric", "numeric", 
                                    "numeric", "numeric", "numeric", 
                                    "numeric", "numeric", "numeric", 
                                    "numeric", "numeric", "numeric", 
                                    "numeric", "numeric", "numeric", 
                                    "numeric", "numeric", "numeric", 
                                    "text", "numeric"))
View(BDD_PEV)

plot(BDD_PEV$BCG)
BCG=ts(BDD_PEV$BCG,start = c(2019,01,01),end =c (2020,10,30),frequency = 12)
plot(BCG)
OPV_3=ts(BDD_PEV$OPV_3,start = c(2019,01),end =c (2020,10),frequency = 12)
OPV_0=ts(BDD_PEV$OPV_0,start = c(2019,01),end =c (2020,10),frequency = 12)
OPV_1=ts(BDD_PEV$OPV_1,start = c(2019,01),end =c (2020,10),frequency = 12)
OPV_2=ts(BDD_PEV$OPV_2,start = c(2019,01),end =c (2020,10),frequency = 12)
IPV=ts(BDD_PEV$IPV,start = c(2019,01),end =c (2020,10),frequency = 12)
DPT1=ts(BDD_PEV$DPT1,start = c(2019,01),end =c (2020,10),frequency = 12)
DPT2=ts(BDD_PEV$DPT2,start = c(2019,01),end =c (2020,10),frequency = 12)
DPT3=ts(BDD_PEV$DPT3,start = c(2019,01),end =c (2020,10),frequency = 12)
MMR=ts(BDD_PEV$MMR,start = c(2019,01),end =c (2020,10),frequency = 12)
AAV=ts(BDD_PEV$AAV,start = c(2019,01),end =c (2020,10),frequency = 12)
EnfComp=ts(BDD_PEV$Enf.Comp,start = c(2019,01),end =c (2020,10),frequency = 12)
Td1=ts(BDD_PEV$Td1,start = c(2019,01),end =c (2020,10),frequency = 12)
Td2=ts(BDD_PEV$Td2,start = c(2019,01),end =c (2020,10),frequency = 12)
Covid=ts(BDD_PEV$Covid,start = c(2019,01),end =c (2020,10),frequency = 12)
BDD_PEV$time_covid=factor(BDD_PEV$time_covid,levels = c(0,1))

BDD_PEV$Period=as.Date(BDD_PEV$Period)

z=cbind(OPV_0,OPV_2,OPV_1,OPV_3,IPV,DPT1,DPT2,DPT3,MMR,AAV,Td1,Td2)

library("gtsummary", lib.loc="/Library/Frameworks/R.framework/Versions/4.0/Resources/library")
BDD_PEV%>%
  tbl_summary()%>%
  add_n()%>%
  bold_labels()

BDD_PEV%>%
  tbl_summary(by = "Period_covid" )%>%
  add_n()%>%
#  add_p()%>%
  bold_labels()

par(mfrow=c(3,3))

plot(cbind(z[,1]),ylab = "OPV_0",xlab="Month per year",pch = 18, col = "black", lty = 1, lwd = 2)
axis(2, at = time(cbind(z[,1])), labels = F)
abline(v=2020.3, col="purple")

plot(cbind(z[,2]),ylab = "OPV_2",xlab="Month per year",pch = 18, col = "black", type = "b", lty = 1, lwd = 1)
axis(2, at = time(cbind(z[,2])), labels = F)
abline(v=2020.3, col="purple")

plot(cbind(z[,3]),ylab = "OPV_1",xlab="Month per year",pch = 18, col = "black", type = "b", lty = 1, lwd = 1)
axis(2, at = time(cbind(z[,3])), labels = F)
abline(v=2020.3, col="purple")

plot(cbind(z[,4]),ylab = "OPV_3",xlab="Month per year",pch = 18, col = "black", type = "b", lty = 1, lwd = 1)
axis(2, at = time(cbind(z[,4])), labels = F)
abline(v=2020.3, col="purple")

plot(cbind(z[,5]),ylab = "IPV",xlab="Month per year",pch = 18, col = "black", type = "b", lty = 1, lwd = 1)
axis(2, at = time(cbind(z[,5])), labels = F)
abline(v=2020.3, col="purple")

plot(cbind(z[,5]),ylab = "IPV",xlab="Month per year",pch = 18, col = "black", type = "b", lty = 1, lwd = 1)
axis(2, at = time(cbind(z[,5])), labels = F)
abline(v=2020.3, col="purple")

plot(cbind(z[,6]),ylab = "DPT1",xlab="Month per year",pch = 18, col = "black", type = "b", lty = 1, lwd = 1)
axis(2, at = time(cbind(z[,5])), labels = F)
abline(v=2020.3, col="purple")

plot(cbind(z[,6]),ylab = "DPT2",xlab="Month per year",pch = 18, col = "black", type = "b", lty = 1, lwd = 1)
axis(2, at = time(cbind(z[,5])), labels = F)
abline(v=2020.3, col="purple")

plot(cbind(z[,6]),ylab = "DPT3",xlab="Month per year",pch = 18, col = "black", type = "b", lty = 1, lwd = 1)
axis(2, at = time(cbind(z[,5])), labels = F)
abline(v=2020.3, col="purple")


ccf(diff(OPV_0),diff(Covid),main="TB cases all form notified AND Ebola")
ccf(diff(OPV_1),diff(Covid),main="TB cases all form notified AND Ebola")

Period_covid=factor(BDD_PEV$Period_covid)


library(ggplot2)
library(ggpubr)
theme_set(theme_pubr())

# Naissance a 6ieme semaine

g = ggplot(BDD_PEV,aes(x=Period,y=BCG)) + geom_point() + geom_smooth(method = "lm", se=F,aes(group=Period_covid)) + theme_bw() +  labs(colour="red") + theme(plot.caption = element_text(lineheight=.5))+geom_vline(xintercept = as.numeric(BDD_PEV$Period[15]), linetype="dashed") + scale_x_date(limit=c(as.Date("2019-01-01"),as.Date("2020-10-30")))

g

c = ggplot(BDD_PEV,aes(x=Period,y=OPV_0)) + geom_point() + geom_smooth(method = "lm", se=F,aes(group=Period_covid)) + theme_bw() +  labs(colour="red") + theme(plot.caption = element_text(lineheight=.5))+ scale_x_date(limit=c(as.Date("2019-01-01"),as.Date("2020-10-30")))+geom_vline(xintercept = as.numeric(BDD_PEV$Period[15]), linetype="dashed")

t= ggplot(BDD_PEV,aes(x=Period,y=DPT1)) + geom_point() + geom_smooth(method = "lm", se=F,aes(group=Period_covid)) + theme_bw() +  labs(colour="red") + theme(plot.caption = element_text(lineheight=.5))+ scale_x_date(limit=c(as.Date("2019-01-01"),as.Date("2020-10-30")))+geom_vline(xintercept = as.numeric(BDD_PEV$Period[15]), linetype="dashed")

s= ggplot(BDD_PEV,aes(x=Period,y=OPV_1)) + geom_point() + geom_smooth(method = "lm", se=F,aes(group=Period_covid)) + theme_bw() +  labs(colour="red") + theme(plot.caption = element_text(lineheight=.5))+ scale_x_date(limit=c(as.Date("2019-01-01"),as.Date("2020-10-30")))+geom_vline(xintercept = as.numeric(BDD_PEV$Period[15]), linetype="dashed")

figure <- ggarrange(c, g,t,s, labels = c("A", "B","D","E"), ncol = 2, nrow = 2)
figure

# 10 a 14 ieme semaine


g = ggplot(BDD_PEV,aes(x=Period,y=DPT2)) + geom_point() + geom_smooth(method = "lm", se=F,aes(group=Period_covid)) + theme_bw() +  labs(colour="red") + theme(plot.caption = element_text(lineheight=.5))+ scale_x_date(limit=c(as.Date("2019-01-01"),as.Date("2020-10-30")))+geom_vline(xintercept = as.numeric(BDD_PEV$Period[15]), linetype="dashed")

c = ggplot(BDD_PEV,aes(x=Period,y=OPV_2)) + geom_point() + geom_smooth(method = "lm", se=F,aes(group=Period_covid)) + theme_bw() +  labs(colour="red") + theme(plot.caption = element_text(lineheight=.5))+ scale_x_date(limit=c(as.Date("2019-01-01"),as.Date("2020-10-30")))+geom_vline(xintercept = as.numeric(BDD_PEV$Period[15]), linetype="dashed")


 t=ggplot(BDD_PEV,aes(x=Period,y=DPT3)) + geom_point() + geom_smooth(method = "lm", se=F,aes(group=Period_covid)) + theme_bw() +  labs(colour="red") + theme(plot.caption = element_text(lineheight=.5))+ scale_x_date(limit=c(as.Date("2019-01-01"),as.Date("2020-10-30")))+geom_vline(xintercept = as.numeric(BDD_PEV$Period[15]), linetype="dashed")

s= ggplot(BDD_PEV,aes(x=Period,y=OPV_3)) + geom_point() + geom_smooth(method = "lm", se=F,aes(group=Period_covid)) + theme_bw() +  labs(colour="red") + theme(plot.caption = element_text(lineheight=.5))+ scale_x_date(limit=c(as.Date("2019-01-01"),as.Date("2020-10-30")))+geom_vline(xintercept = as.numeric(BDD_PEV$Period[15]), linetype="dashed")

w= ggplot(BDD_PEV,aes(x=Period,y=IPV)) + geom_point() + geom_smooth(method = "lm", se=F,aes(group=Period_covid)) + theme_bw() +  labs(colour="red") + theme(plot.caption = element_text(lineheight=.5))+ scale_x_date(limit=c(as.Date("2019-01-01"),as.Date("2020-10-30")))+geom_vline(xintercept = as.numeric(BDD_PEV$Period[15]), linetype="dashed")

f= ggplot(BDD_PEV,aes(x=Period,y=EnfComp)) + geom_point() + geom_smooth(method = "lm", se=F,aes(group=Period_covid)) + theme_bw() +  labs(colour="red") + theme(plot.caption = element_text(lineheight=.5))+ scale_x_date(limit=c(as.Date("2019-01-01"),as.Date("2020-10-30")))+geom_vline(xintercept = as.numeric(BDD_PEV$Period[15]), linetype="dashed")

figure <- ggarrange(c, g,t,s,w,f, labels = c("A", "B","D","E","F","G"), ncol = 2, nrow = 3)
figure

# AVT

wl= ggplot(BDD_PEV,aes(x=Period,y=Td1)) + geom_point() + geom_smooth(method = "lm", se=F,aes(group=Period_covid)) + theme_bw() +  labs(colour="red") + theme(plot.caption = element_text(lineheight=.5))+ scale_x_date(limit=c(as.Date("2019-01-01"),as.Date("2020-10-30")))+geom_vline(xintercept = as.numeric(BDD_PEV$Period[15]), linetype="dashed")

cf= ggplot(BDD_PEV,aes(x=Period,y=Td2)) + geom_point() + geom_smooth(method = "lm", se=F,aes(group=Period_covid)) + theme_bw() +  labs(colour="red") + theme(plot.caption = element_text(lineheight=.5))+ scale_x_date(limit=c(as.Date("2019-01-01"),as.Date("2020-10-30")))+geom_vline(xintercept = as.numeric(BDD_PEV$Period[15]), linetype="dashed")

figure <- ggarrange(wl,cf, labels = c("A", "B"), ncol = 1, nrow = 2)
figure


# analyse des series temporelles interronpues

library(its.analysis)
# BCG
year <-BDD_PEV$year
depv <-BDD_PEV$BCG
interruption <- as.factor(BDD_PEV$time_covid)
cov1 <- BDD_PEV$MMR_BCG

x <- as.data.frame(cbind(year, depv, interruption, cov1))

itsa.model(data=x, time="year", depMMR="depv", interrupt_MMR = "interruption",
           alpha=0.05, bootstrap=TRUE, Reps = 10)

# OPV
year <-BDD_PEV$year
depv <-BDD_PEV$OPV_0
interruption <- as.factor(BDD_PEV$time_covid)
cov1 <- BDD_PEV$OPV_0

x <- as.data.frame(cbind(year, depv, interruption, cov1))

itsa.model(data=x, time="year", depMMR="depv", interrupt_MMR = "interruption",
           alpha=0.05, bootstrap=TRUE, Reps = 10)


# OPV_1
year <-BDD_PEV$year
depv <-BDD_PEV$OPV_1
interruption <- as.factor(BDD_PEV$time_covid)
cov1 <- BDD_PEV$OPV_1

x <- as.data.frame(cbind(year, depv, interruption, cov1))

itsa.model(data=x, time="year", depMMR="depv", interrupt_MMR = "interruption",
           alpha=0.05, bootstrap=TRUE, Reps = 10)


# OPV_2
year <-BDD_PEV$year
depv <-BDD_PEV$OPV_2
interruption <- as.factor(BDD_PEV$time_covid)
cov1 <- BDD_PEV$OPV_2

x <- as.data.frame(cbind(year, depv, interruption, cov1))

itsa.model(data=x, time="year", depMMR="depv", interrupt_MMR = "interruption",
           alpha=0.05, bootstrap=TRUE, Reps = 10)

# OPV_3

year <-BDD_PEV$year
depv <-BDD_PEV$OPV_3
interruption <- as.factor(BDD_PEV$time_covid)
cov1 <- BDD_PEV$OPV_3

x <- as.data.frame(cbind(year, depv, interruption, cov1))

itsa.model(data=x, time="year", depMMR="depv", interrupt_MMR = "interruption",
           alpha=0.05, bootstrap=TRUE, Reps = 10)

# IPV

year <-BDD_PEV$year
depv <-BDD_PEV$IPV
interruption <- as.factor(BDD_PEV$time_covid)
cov1 <- BDD_PEV$IPV

x <- as.data.frame(cbind(year, depv, interruption, cov1))

itsa.model(data=x, time="year", depMMR="depv", interrupt_MMR = "interruption",
           alpha=0.05, bootstrap=TRUE, Reps = 10)


# DPT1

year <-BDD_PEV$year
depv <-BDD_PEV$DPT1
interruption <- as.factor(BDD_PEV$time_covid)
cov1 <- BDD_PEV$DPT1

x <- as.data.frame(cbind(year, depv, interruption, cov1))

itsa.model(data=x, time="year", depMMR="depv", interrupt_MMR = "interruption",
           alpha=0.05, bootstrap=TRUE, Reps = 10)

# DPT2

year <-BDD_PEV$year
depv <-BDD_PEV$DPT2
interruption <- as.factor(BDD_PEV$time_covid)
cov1 <- BDD_PEV$DPT2

x <- as.data.frame(cbind(year, depv, interruption, cov1))

itsa.model(data=x, time="year", depMMR="depv", interrupt_MMR = "interruption",
           alpha=0.05, bootstrap=TRUE, Reps = 10)

# DPT3

year <-BDD_PEV$year
depv <-BDD_PEV$DPT3
interruption <- as.factor(BDD_PEV$time_covid)
cov1 <- BDD_PEV$DPT3

x <- as.data.frame(cbind(year, depv, interruption, cov1))

itsa.model(data=x, time="year", depMMR="depv", interrupt_MMR = "interruption",
           alpha=0.05, bootstrap=TRUE, Reps = 10)


# MMR

year <-BDD_PEV$year
depv <-BDD_PEV$MMR
interruption <- as.factor(BDD_PEV$time_covid)
cov1 <- BDD_PEV$MMR

x <- as.data.frame(cbind(year, depv, interruption, cov1))

itsa.model(data=x, time="year", depMMR="depv", interrupt_MMR = "interruption",
           alpha=0.05, bootstrap=TRUE, Reps = 10)


# library(readxl)
# nobre_cas_RR <- read_excel("~/Downloads/nobre cas RR à ce jour 14-8-2020 (1) (1).xlsx")
# View(nobre_cas_RR) 
# 
# g = ggplot(nobre_cas_RR,aes(x=Date,y=Cas_TBMR)) + geom_point() + geom_smooth(method = "lm", se=F)+theme_bw() +  labs(colour="red") + theme(plot.caption = element_text(lineheight=.5))+geom_vline(xintercept = as.numeric(nobre_cas_RR$Date[16]), linetype="dashed") #+ scale_x_date(limit=c(as.Date("2019-01-01"),as.Date("2020-06-30")))
# g
# 
# 
# Cas_TBMR=ts(nobre_cas_RR$Cas_TBMR,start = c(2019,01),end =c (2020,7),frequency = 12)
# plot(Cas_TBMR)
# axis(2,  labels = F)
# abline(v=2020.3, col="purple")
# 
# 
# year <-nobre_cas_RR$Date
# depv <-nobre_cas_RR$Cas_TBMR
# interruption <- as.factor(nobre_cas_RR$time)
# cov1 <- nobre_cas_RR$MMR
# 
# x <- as.data.frame(cbind(year, depv, interruption, cov1))
# 
# itsa.model(data=x, time="year", depMMR="depv", interrupt_MMR = "interruption",
#            alpha=0.05, bootstrap=TRUE, Reps = 10)
```
